# SARS-CoV-2 antibodies in the Southern Region of New Zealand, 2020

**DOI:** 10.1101/2020.10.20.20215616

**Authors:** Alyson Craigie, Reuben McGregor, Alana Whitcombe, Lauren Carlton, David Harte, Michelle Sutherland, Matthew Parry, Erasmus Smit, Gary McAuliffe, James Ussher, Nicole Moreland, Susan Jack, Arlo Upton

## Abstract

**Background:** During New Zealand’s first outbreak in early 2020 the Southern Region had the highest per capita SARS-CoV-2 infection rate. PCR testing was initially limited by a narrow case definition and limited laboratory capacity, so cases may have been missed.

**Objectives:** To evaluate the Abbott^®^ SARS-CoV-2 IgG nucleocapsid assay, alongside spike-based assays, and to determine the frequency of antibodies among PCR-confirmed and probable cases, contacts, and higher risk individuals in the Southern Region of NZ.

**Study design:** Pre-pandemic sera (n=300) were used to establish assay specificity and sera from PCR-confirmed SARS-CoV-2 patients (n=78) to establish sensitivity. For prevalence analysis, all samples (n=1214) were tested on the Abbott assay, and all PCR-confirmed cases (n=78), probable cases (n=9), and higher risk individuals with *grey-zone* (n=14) or positive results (n=11) were tested on four additional SARS-CoV-2 serological assays.

**Results:** The median time from infection onset to serum collection for PCR-confirmed cases was 14 weeks (range 11-17 weeks). The Abbott assay demonstrated a specificity of 99.7% (95% CI, 98.2%-99.99%) and a sensitivity of 76.9% (95% CI, 66.0%-85.7%). Spike-based assays demonstrated superior sensitivity ranging 89.7-94.9%. Nine previously undiagnosed sero-positive individuals were identified, and all had epidemiological risk factors.

**Conclusions:** Spike-based assays demonstrated higher sensitivity than the Abbott IgG assay, likely due to temporal differences in antibody persistence. No unexpected SARS-CoV-2 infections were found in the Southern region of NZ, supporting the elimination status of the country at the time this study was conducted.

## Introduction

Severe Acute Respiratory Syndrome Coronavirus 2 (SARS-CoV-2) and the disease it causes, COVID-19, were first detected in Wuhan, China in December 2019 [1]. By 22 May 2020, there were nearly 5 million confirmed COVID-19 cases worldwide, with ∼336000 deaths [2]. During New Zealand’s (NZ) first outbreak (28 February to 22 May 2020) a total of 1154 PCR-confirmed and 350 probable cases were identified, with 22 COVID-19 related deaths [3].

This serological study is focused on the Southern District Health Board (SDHB) region in NZ, which had the largest number of COVID-19 cases per capita during NZ’s first outbreak (216 total cases; ∼66/100,000 population), significantly higher than the national average (∼30/100,000) [3]. This region also includes the tourism hub of Queenstown, where community transmission took place. PCR testing for SARS-CoV-2 was initially restricted in NZ due to a narrow case definition and limited access to diagnostic reagents.

Reverse transcription polymerase chain reaction (henceforth referred to as PCR) from a nasopharyngeal and/or oropharyngeal swab or lower respiratory tract sample is the gold standard method for detecting acute SARS-CoV-2 infection [4], whereas serological tests can provide information on past infection [4]. The majority of SARS-CoV-2 serological assays detect antibodies against spike (S) and/or nucleocapsid (N) proteins [4]. Differing degrees of sequence conservation between N and S proteins (including S1 and receptor binding domains (RBD)), and homologous proteins from other coronaviruses, together with differences in the magnitude and kinetics of antibody responses to these antigens may impact assay performance [5]. Several serological assays are now commercially available, including for use on high-throughput, random access analysers such as the Abbott Architect.

The aims of this study were threefold. Firstly, to investigate the sensitivity and specificity of the Abbott Architect SARS-CoV-2 IgG assay, based on the N protein, together with several plate-based assays that utilise the S protein and/or S protein domains. Secondly, to determine the frequency of SARS-CoV-2 IgG antibodies among contacts of cases and other higher risk individuals in the SDHB region to determine whether cases were missed during the outbreak. Thirdly, to assess the likelihood of infection among those diagnosed as ‘probable’ cases.

## Materials and Methods

### Study Protocol

This study was performed at Southern Community Laboratories, in conjunction with the SDHB, WellSouth (a primary healthcare organisation), University of Otago, University of Auckland, and the Institute of Environmental Science and Research. It was approved by the Health and Disability Ethics Committee (20/NTB/101).

PCR-confirmed cases, probable cases (PCR test negative or not performed, but classified as a case based on their exposure history (household contact) and clinical symptoms [3]), and their contacts were contacted for recruitment via the local public health unit. The remaining higher risk individuals (frontline healthcare workers, tourism workers, or Queenstown residents) were recruited via posters and media (print, television, and social media). Participants completed a REDCap online survey [6]. The questionnaire included demographic details and questions about which higher risk category they associated with, if they had any contact with known COVID-19 cases, and if they recalled having symptoms consistent with COVID-19 before or during the lockdown period (alert level three and four, between 23^rd^ March to May 12^th^ 2020) (Table 1). Blood samples were collected between 4^th^ June and 4^th^ August 2020. The severity of COVID-19 in the PCR-confirmed group was classified on the basis of their symptoms and the level of hospital care provided: 1 – asymptomatic to mild cold-like symptoms (n=35); 2 – moderate: cough, fever and chills (n=39); 3 – moderately severe: admitted for assessment (n=3); 4 – severe: admitted and given supplemental oxygen therapy (n=1); 5 – critical: admitted to ICU (n=0).

**Table 1.** Patient demographics.

|  |  | **Total** | **PCR-confirmed cases** | **Probable cases** | **Higher risk group** |
| --- | --- | --- | --- | --- | --- |
| **Total** |  | 1214<br>(100%) | 78<br>(6%) | 9<br>(1%) | 1127<br>(93%) |
| **Age (years)** | Median | 46 | 51 | 49 | 46 |
|  | Range | 4-90 | 17-81 | 10-59 | 4-90 |
| **Gender** | M/F | 306/908 | 32/46 | 3/6 | 271/856 |
| **Higher-risk group category** | Household contacts of known or probable cases |  |  |  | 40 (4%) |
|  | Non-household contacts of known or probable cases |  |  |  | 276 (25%) |
|  | Frontline healthcare workers |  |  |  | 702 (62%) |
|  | Tourism worker |  |  |  | 60 (5%) |
|  | Queenstown resident |  |  |  | 208 (19%) |
|  | One or more COVID-19 symptoms reported |  |  |  | 631 (56%) |

To determine assay specificity, 300 de-identified antenatal sera collected from early-mid 2019 (pre-pandemic), were used (after being stored at -20°C for up to 12 months).

### Serological assays

The assays utilised are summarised in Table 2. The primary assay was the Abbott Architect SARS-CoV-2 IgG chemiluminescent microparticle immunoassay (CMIA) (Abbott, Chicago, USA). Samples were analysed on the Abbott Architect i2000SR Immunoassay Analyzer following manufacturer’s instructions.

**Table 2.** Summary of the investigated SARS-CoV-2 assays

| Assay | SARS-CoV-2 antigen target | Company | Positivity threshold | Platform | Sensitivity (according to manufacturer) | Specificity |
| --- | --- | --- | --- | --- | --- | --- |
| Abbott Architect SARS-CoV-2 IgG | N protein | Abbott, Chicago, Illinois, USA | $\geq 1.40$ S/C | Abbott Architect (CMIA) | 0 -100% (day 0 to $\geq 14$ days after disease onset) | 99.6% |
| *In-house* SARS-CoV-2 two-stage IgG ELISA | RBD/S protein | *In-house* | RBD: $\geq 0.2$ OD<br>Spike: $\geq 300$ titre | Manual ELISA | n/a | n/a |
| Wantai SARS-CoV-2 total antibody ELISA | RBD/S protein | Beijing Wantai Biological Pharmacy, Beijing, China | $\geq 1$ A/C.O. | Manual ELISA | 94.5 % (dependent on specimen collection time and time of disease onset) | 100% |
| Euroimmun Anti-SARS-CoV-2 ELISA (IgG) | S1 protein | Euroimmun, Lubeck, Germany | $\geq 1.1$ ratio | Manual ELISA | $\leq 10$ days = 60.2%<br>>10 days = 98.6% | 92.0% |
| cPass® sVNT | S1 protein | GenScript, New Jersey, USA | $\geq 20$ % inhibition | Manual sVNT | 95.0% | 100% |

N: Nucleocapsid; RBD: Receptor Binding Domain; S: Spike; CMIA: Chemiluminescent microparticle immunoassay; sVNT: Surrogate Virus Neutralisation Test

The *in-house* two-step ELISA was adapted from published protocols [7] as described [8]. In step one serum diluted 1:100 was screened against RBD, with IgG binding detected with a peroxidase-labelled anti-human IgG secondary. Samples with an optical density (OD, 450-570nm) above the cut-off (>0.2) were titrated in a 3-fold dilution series against the S protein in step two and considered positive if OD>0.2 in at least two consecutive wells in the confirmatory S protein ELISA.

The Wantai SARS-CoV-2 ELISA (Beijing Wantai Biological Pharmacy Enterprise, Beijing, China) and EUROIMMUN SARS-CoV-2 IgG ELISA (EUROIMMUN AG, Lübeck, Germany) were performed according to the manufacturer’s instructions. The cPass® surrogate viral neutralisation test (sVNT) (GenScript, New Jersey, USA) measures the presence of neutralising antibodies (NAbs) that are capable of blocking the interaction between RBD and hACE2 [9] and was performed according to manufacturer’s instructions. Samples with percentage inhibition ≥20% were defined as having neutralising antibodies.

To assess cross-reactivity of antenatal sera with other human coronaviruses (HCoV), ELISA were performed using S1 antigens from the HKU1, NL63, and SARS-CoV-2 (Sino Biological, Beijing, China) at 1:300 sera dilution as described [8].

### Testing protocol

The antenatal samples and serum from PCR-confirmed and probable cases were tested on all five assays. All sera from the higher risk group (n=1127) were tested on the Abbott assay. Samples from the higher risk group that classified as positive (≥1.4 S/C), or as negative but ≥ 0.5 S/C (i.e. 0.5–1.39 S/C, defined as the *grey-zone* on the basis of a receiver operating characteristic (ROC) curve) were tested on all assays (n=25).

### Statistical analysis

Statistical analysis was performed using Prism 8 (GraphPad) or R (version 3.6.3) within R Studio (version 1.2.5033); a *P*-value of ≤0.05 was considered statistically significant. To assess sensitivity, we chose the PCR results as gold standard. Equivocal results were considered negative for the statistical analyses. For the higher risk group, true sero-positivity was defined by positivity in two or more of the five assays. False positivity was defined as positivity in only one of the five assays.

## Results

### Participant characteristics

1214 individuals gave informed consent and participated in the study. Of these, 78 were PCR-confirmed cases, nine were probable cases, and 1127 were in the higher risk group. The median age of the mostly female (75%) participants was 46 years (range 4– 90 years) (Table 1). Of the 1127 higher risk participants, 37% had had a negative PCR test, 62% self-identified as frontline healthcare workers in the southern region, and 56% reported one or more symptoms in the two weeks prior to and during the February-May COVID-19 outbreak. For the PCR-confirmed and probable cases, median time from symptom onset to blood collection was 14 weeks (range 11-17 weeks).

### Assay performance

Overall performance of the assays is summarised in Table 3. Specificity was high across all assays ranging from 99.3% (95% CI, 97.6-99.9%) to 100% (95% CI, 98.8-100.0%). The antenatal sera used to determine specificity showed broad reactivity with S1 protein antigens from HCoV (HKU1 and NL63), but not SARS-CoV-2 (Supplementary figure 1). Sensitivity ranged from 76.9% (95% CI: 66.0-85.7%) for the Abbott assay, to 94.9% (95% CI, 87.4-98.6%) for the Wantai assay (Figure 1, Table 3). Eighteen (23.1%) PCR-confirmed cases tested negative on the Abbott; raw values ranged from 0.14-1.39 S/C. Eleven of these were positive on ≥3 other assays, four were positive on two other assays, one was positive on one other assay, and two were negative on all other assays.

**Table 3.** Sensitivity and specificity of the investigated SARS-CoV-2 assays

| Assay | SARS-CoV-2 antigen | Sensitivity (%) | Specificity (%) |
| --- | --- | --- | --- |
| Abbott Architect SARS-CoV-2 IgG<br>(using manufacturer cut-off of $\geq 1.40$) | N protein | 76.9 (60/78)<br>(95% CI: 66.0-85.7) | 99.7 (299/300)<br>(95% CI: 98.2-99.99) |
| Abbott Architect SARS-CoV-2 IgG<br>(using revised cut-off of $\geq 0.50$) | N protein | 94.9 (74/78)<br>(95% CI: 87.4-98.6) | 98.3 (295/300)<br>(95% CI: 96.2-99.5) |
| *In-house* SARS-CoV-2 two-stage IgG ELISA | RBD/S protein | 91.0 (71/78)<br>(95% CI: 82.4-96.3) | 100 (300/300)<br>(95% CI: 98.8-100.0) |
| Wantai SARS-CoV-2 total antibody ELISA | RBD/S protein | 94.9 (74/78)<br>(95% CI: 87.4-98.6) | 99.3% (298/300)<br>(95% CI: 97.6-99.9) |
| Euroimmun Anti-SARS-CoV-2 ELISA (IgG)* | S1 protein | 89.7 (70/78)<br>(95% CI: 80.8-95.5) | 100 (300/300)<br>(95% CI: 98.8-100.0) |
| cPass® sVNT | Neutralising antibodies | 88.5% (69/78)<br>(95% CI: 79.2-94.6%) | 100% (300/300)<br>(95% CI: 98.8-100.0%) |

\*Equivocal results considered negative

**Figure 1.**
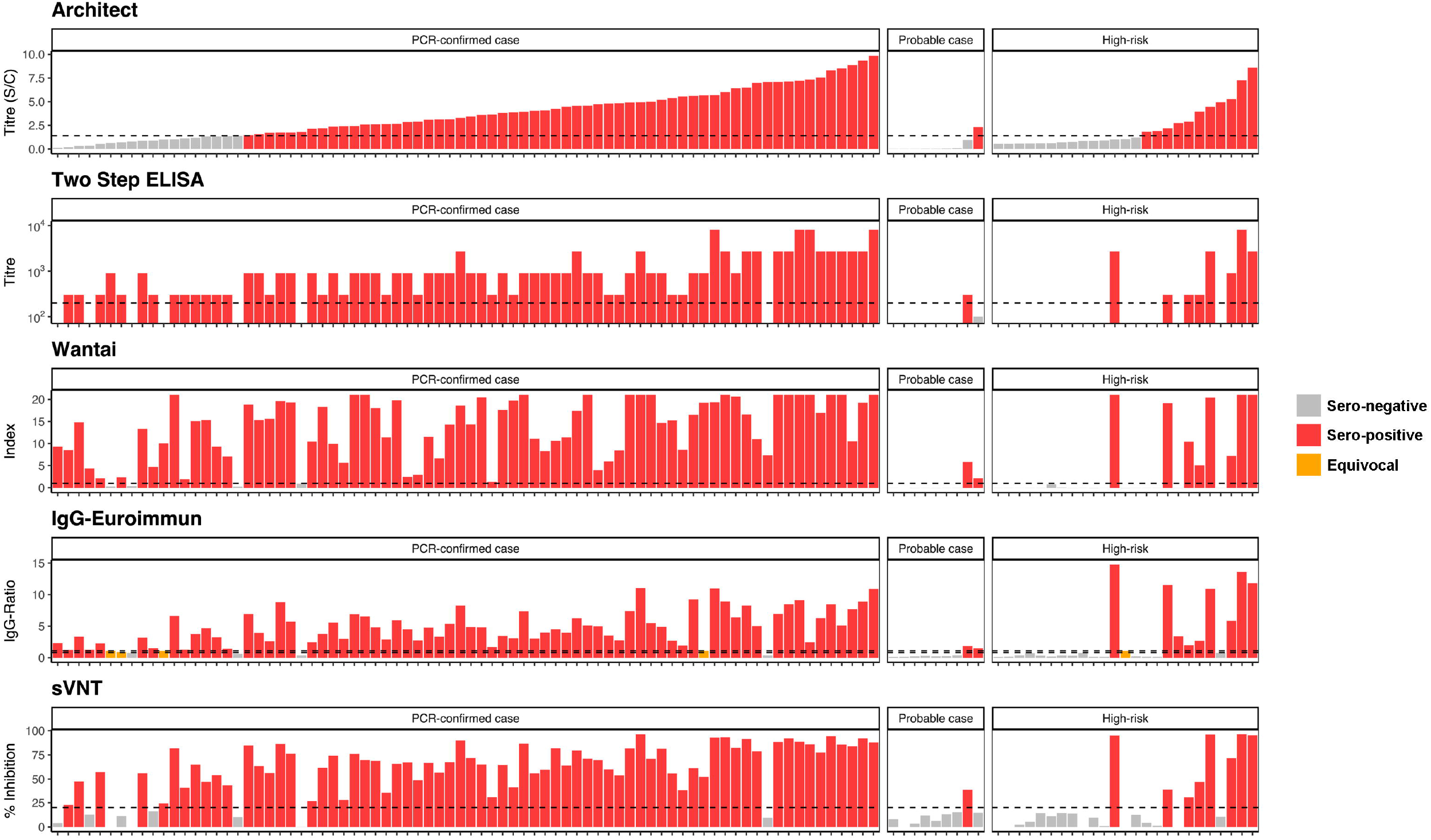
Antibody levels for the examined assays for the samples tested on all five assays (all PCR-confirmed cases, all probable cases, and higher risk samples in the *grey-zone* (0.5-1.39 S/C) or positive (≥1.4 S/C) results on the Abbott assay) (n=112). Dashed horizontal lines show assay specific cut-off.

The sensitivity of the Abbott assay was unexpectedly low and prompted a ROC analysis that showed a cut-off of 0.55 S/C could achieve much greater sensitivity (93.6%) without a significant loss in specificity (98.7%) (Supplementary figure 2). A *grey-zone* approach was therefore utilised for analysis of the higher risk and probable cases to rule out potential false negatives: samples with 0.5–1.39 S/C were measured on the other four assays.

### Neutralising anti-SARS-CoV-2 antibodies

The sVNT assay was used to measure NAbs. For the PCR-confirmed group, 88.5% (69/78) had detectable NAbs. When the PCR-confirmed patients were stratified by disease severity, there was a small but significant increase in the level of NAbs in those with more severe disease (*P* < 0.05) (Supplementary figure 3).

### Antibody detection among higher risk individuals

Eleven individuals of the higher risk group (0.98%) had positive results on the Abbott assay (Figure 1). Eight of these were also positive on ≥1 other assays, indicating true sero-positivity. Three Abbott positive results were considered false positives as they were negative on all four other assays. There were fourteen Abbott results that fell in the *grey-zone* (0.5 – 1.39 S/C). Thirteen (93%) were negative on all other assays and classified as sero-negative. One individual was positive on all four other assays and considered sero-positive.

Thus, in total we detected nine additional possible COVID-19 infections: one was a PCR-confirmed case diagnosed outside of the Southern region; six had consistent travel history (Western Europe/UK) and symptoms; and two were close contacts of PCR-confirmed cases reporting consistent symptoms.

### Antibody detection among probable cases

Of the nine probable cases, one was positive on four of five assays while another was positive on three of five assays, suggesting likely infection (Figure 1). The remaining seven were negative by all assays, and the Abbott assay raw values of these ranged from 0.01 – 0.04 S/C, suggesting that these were unlikely to have had COVID-19 infection.

## Discussion

Using a cohort of PCR-confirmed cases to assess sensitivity, we found suboptimal performance of the Abbott assay at 11-17 weeks post infection with a sensitivity of 76.9%, lower than previously published data [10-12] and manufacturer’s claim (100% after 14 days). It is likely two factors contributed to this. Firstly, most of the cases in the SDHB region were not hospitalised, and there is evidence that antibody levels correlate with disease severity [13]. Secondly, a median of 14 weeks (range 11-17 weeks) had lapsed between symptom onset and serum collection; N protein antibodies are reported to decline relatively quickly post-infection [14]. In contrast, the sensitivity of plate-based assays based on S protein was higher (89.7-94.9%), which is in keeping with reports that antibodies against S protein persist for longer than those to N protein [14].

In our hands, the Abbott assay specificity was 99.7% (95% CI, 98.2%-99.99%), comparable to the manufacturer’s claim (99.6%). However, given the very low prevalence of COVID-19 infection in NZ, the positive-predictive value will be relatively low. Thus, we suggest an orthogonal testing algorithm (a second assay, using a different target) as a supplemental assay before reporting results as true positives.

S protein is the main target for SARS-CoV-2 NAbs [15]. In this study NAbs were measured using a sVNT [9] with 88.5% of the PCR-confirmed cases having detectable NAbs 11-17 weeks post-infection, with lower NAbs among those with mild symptoms. A decline in NAb levels has been noted in recent reports [13], but further studies are needed to fully understand these immunokinetics and implications for protection against reinfection.

This study of 1127 individuals who self-identified as being higher risk for SARS-CoV-2 infection identified a further nine infections (0.8%). Of these, all had epidemiological risks including travel to Europe during their outbreak and/or being a close contact of a known case. Undiagnosed infection was not detected among front-line healthcare workers, tourism workers, and casual contacts of known cases. It is possible that cases of infection may have been missed as the Abbott assay, which was used as our initial screening assay, demonstrated sub-optimal sensitivity. However, the *grey-zone* approach utilised, based on a ROC analysis, improved the sensitivity to 94.9%.

Given the imperfect sensitivity, and unknown prevalence among the tested population in our region, it is difficult to estimate the true number of cases of infection that may have been missed. The Rogan-Gladen estimator [16] allows us to estimate the actual prevalence in the higher risk group, considering the uncertainties in the sensitivity and specificity of the test. Using a threshold of 1.4 S/C, the estimated actual prevalence in the higher risk group is 0.8% (95% CI: 0.0-2.0%). Using a threshold of 0.5 S/C, the estimated actual prevalence is 2.0% (95% CI: 0.8-3.2%). To incorporate the effect of secondary orthogonal testing, it is convenient to carry out a Bayesian statistical analysis (see Flor et al [17] for a recent discussion of Bayesian methods in prevalence estimation). Applying the secondary tests to the eleven samples that tested positive with a threshold of 1.4 S/C, the estimated actual prevalence in the higher-risk group is 0.9% (95% credible interval: 0.4-1.7%). Applying the secondary tests to the 25 samples that tested positive with a threshold of 0.5 S/C, the estimated actual prevalence is 0.8% (95% credible interval: 0.4-1.5%). The value of orthogonal testing is two-fold: (i) we obtain more precise estimates of actual prevalence, (ii) the estimates do not appear to depend strongly on the threshold used in the primary test, as evidenced by the concordance of the obtained estimates.

An unexpected finding was that seven of the nine individuals diagnosed with ‘probable’ infection and included in NZ’s official tally were sero-negative despite being tested on all five assays. While acknowledging the delay (approximately three months) in serum collection and the possible impact on sensitivity, it is highly likely that at least some of these individuals did not have infection. This highlights the role of serology in the diagnostic algorithm where PCR is negative despite symptoms and epidemiological risks; further testing of NZ’s remaining 341 probable cases may be warranted.

Our study has some limitations. Firstly, the delay in specimen collection after the outbreak likely had an impact on the Abbott assay sensitivity. We cannot be certain that undiagnosed cases were not missed using the Abbott assay as our screening test. However, every effort was made to mitigate against this by lowering the cut-off for the initial Abbott screening assay. Lastly, it is important to note that this is not a sero-prevalence study. Participants who self-identified as higher risk were actively recruited, therefore the sero-positivity rate calculated in this group in the SDHB region cannot be extrapolated to the general population.

## Conclusion

In conclusion, our study shows that the COVID-19 outbreak in the SDHB region in early 2020 was largely confined to the PCR-confirmed cases and those identified as at higher risk due to recent travel and/or close contact with a known case. We found little evidence of undetected infection among the individuals in the SDHB region who were close contacts identified by Public Health, or whose job or place of residence placed them at higher risk. The N protein based Abbott assay demonstrated the lowest sensitivity of the assays investigated, likely impacted by the delay in serum collection. Whilst this may lead to missed cases, the utility of a high throughput system for large scale testing does, to a degree, offset this significant limitation, especially when combined with secondary S protein assays of higher sensitivity. When designing a SARS-CoV-2 serological assay algorithm, the purpose of testing is a major consideration, with different assay combinations suitable for high-throughput sero-prevalence purposes versus individual level clinical diagnostics.

## Data Availability

The data that support the findings of this study are available from the corresponding author, AU, upon reasonable request.

## Author statement

**Alyson Craigie:** Conceptualisation, Methodology, Formal analysis, Investigation, Data curation, Writing – Original Draft, Visualisation, Project Administration. **Reuben McGregor:** Formal analysis, Investigation, Visualisation. **Alana Whitcombe:** Investigation. **Lauren Carlton:** Investigation. **David Harte:** Investigation. **Michelle Sutherland:** Investigation. **Matthew Parry:** Formal analysis. **Erasmit Smit:** Conceptualisation. **Gary McAuliffe:** Writing – review and editing. **James Ussher:** Conceptualisation, Methodology, Writing – review and editing, Supervision. **Nicole Moreland:** Conceptualisation, Methodology, Writing – review and editing. **Susan Jack:** Conceptualisation, Methodology, Resources, Writing – review and editing. **Arlo Upton:** Conceptualisation, Methodology, Resources Writing – review and editing, Supervision, Project administration.

## Acknowledgements

We thank the team behind the Southern COVID-19 Serology Study for facilitating sample collection, delivery, and processing for the SARS-CoV-2 serological testing. Sample collection, processing, and Abbott Architect consumables were funded by Southern Community Laboratories. This work was also funded in part by the School of Medicine Foundation (University of Auckland) and the COVID-19 Innovation Acceleration Fund (Ministry of Business, Innovation and Employment).

We thank Linfa Wang at Duke-NUS and GenScript for providing sVNT testing kits and technical advice. We are grateful to Drs Campbell Sheen (Callaghan Innovation) and James Dickson (University of Auckland) for providing antigens for the *in-house* ELISA.

