## Supplementary Figures for "SARS-CoV-2 antibodies in the Southern Region of New Zealand, 2020"

**Supplementary figure 1.** Serum IgG reactivity of pre-pandemic antenatal sera with human coronavirus S1 protein. ELISA were performed with sera (n=300) against S1 proteins of one α-HCoV (NL63) and one β- HCoV (HKU1) as well as S1 protein of SARS-CoV-2. One way ANOVA was conducted and determined statistically significant between group means, with a p-value of <0.0001. Post hoc testing using Tukey’s multiple comparison was then conducted, ****p<0.0001.

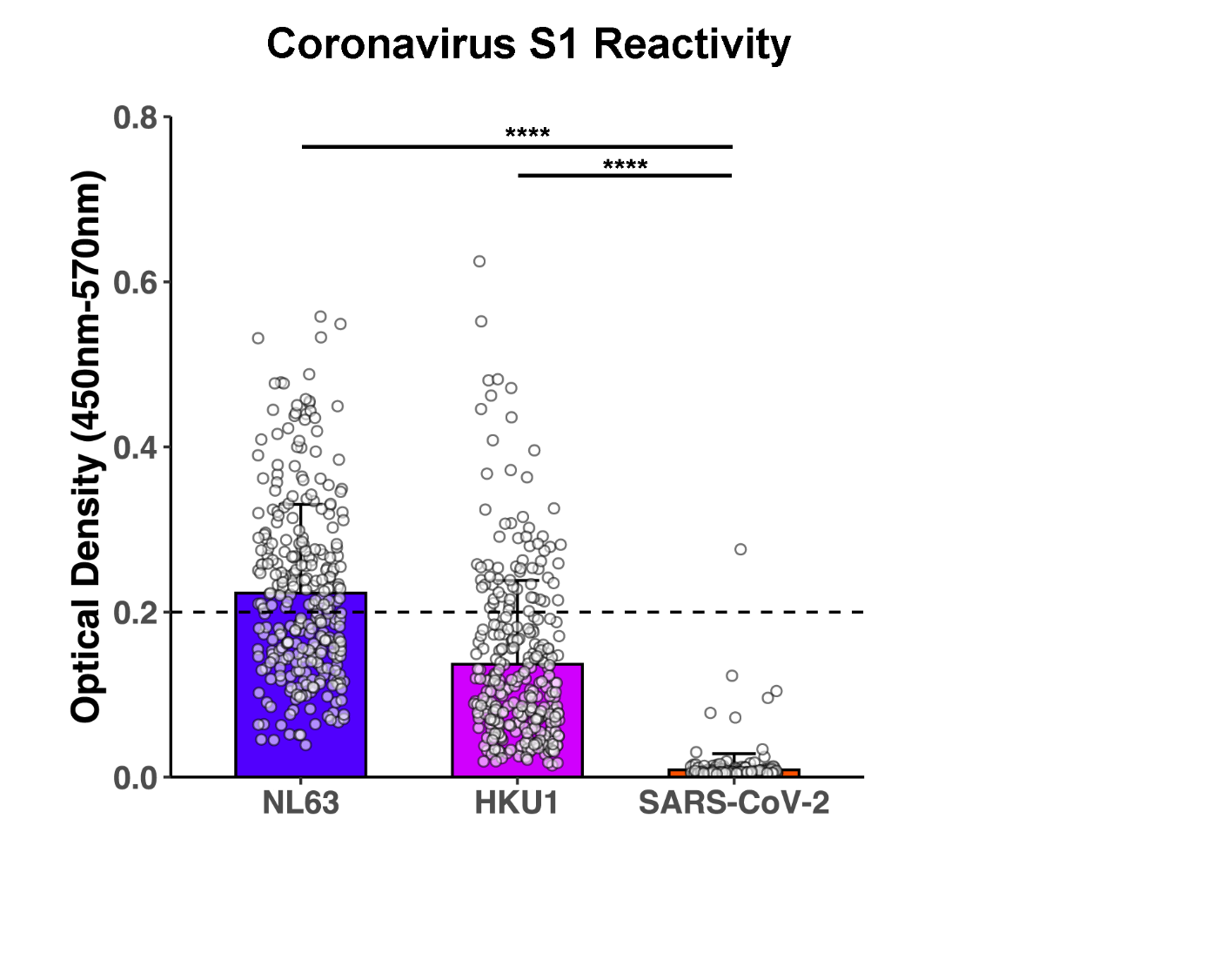

**Supplementary figure 2.** A receiver operating characteristic (ROC) curve, shown in blue, for the Abbott assay derived from the test S/C values of 78 positive samples (PCR-confirmed cases) and 300 negative samples (antenatal serum samples from early- to mid-2019). The location of the manufacturer’s cut-off (1.4 S/C) and the *grey-zone* cut-off (0.5 S/C) are included, showing the large gain in sensitivity for the *grey-zone* cut-off. The line of no-discrimination is shown in grey.

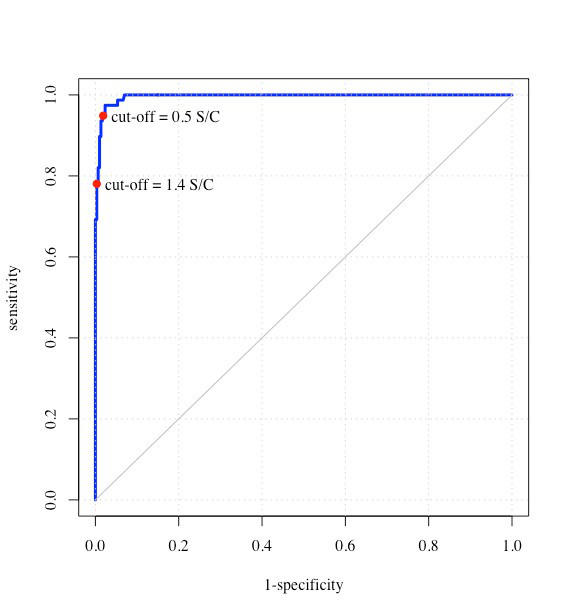

**Supplementary figure 3.** Neutralising antibody levels (% inhibition from sVNT assay) in PCR-confirmed COVID-19 individuals with varying symptom severity. Individuals with mild symptoms (severity level 1) had significantly lower levels of neutralising antibodies than those with moderate to severe symptoms (severity level 2-4). Wilcoxon rank sum test was conducted comparing disease severity of 1 (n=35) and 2-4 (n=43), ***P* <0.05.

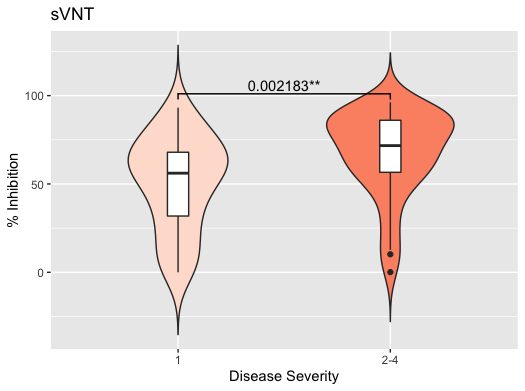
